# LARisk: An R package for lifetime attributable risk from radiation exposure

**DOI:** 10.1101/2022.02.21.22271307

**Authors:** Juhee Lee, Young Min Kim, Yeongwoo Park, Eunjin Jang, Jinkyung Yoo, Songwon Seo, Eun Shil Cha, Won Jin Lee

## Abstract

The lifetime radiation associated risk of cancer incidence has been of interest at least seventy years after atomic bombings of Hiroshima and Nagasaki in 1945. Since most of radiation epidemiological studies are retrospective, it is important to assess the potential magnitude of radiation-related cancer risk. The R-package so-call **LARisk** implements to easily calculate lifetime attributable risks (LAR) of radiation associated cancer incidence presenting bootstrap-based confidence intervals of the estimated risks. The main characteristics of the **LARisk** package are to flexibly compute the LAR-associated risks, effectively summarize large data, and promptly update domain-knowledge such as new regression coefficients for excess relative risk or excess absolute risk models, population information, etc. The manuscript provides a complete guide for usage of the **LARisk** package, which will enable to contribute the prospective studies in radiation epidemiology.

## Introduction

The lifetime radiation related risk of cancer incidence remains elevated for at least seventy years after atomic bombings of Hiroshima and Nagasaki, Japan in 1945 [1]. Estimating radiation related cancer risk has made steady progress [1, 2], but most studies have been retrospective. Forward projection of radiation related cancer risk is essential to allow predictive studies of irradiated groups to ultimately control excess radiation exposure occurring, e.g. from medical radiography (computed tomography, CT) scans [3, 4].

There are several reports to discuss lifetime radiation related cancer risk projection models. The U.S. Nuclear Regulatory Commission developed the NUREG model to predict health effects from radiation exposure at nuclear power plants, considering the dose and dose-rate effectiveness factor (DDREF) and age-specific risk coefficients [5]. The U.S. National Research Council proposed the BEIR VII model [6] for radiation exposed groups, focusing on cancer risk projection from low linear energy transfer (LET) radiation exposure with weighted average estimates from excess relative risk (ERR) and excess absolute risk (EAR) models [6]. The BEIR VII risk models were based primarily on the Life Span Study (LSS) study from the Radiation Effects Research Foundation (RERF), which has advantages of its large population, including of both sexes and all ages, long follow-up, and high-quality mortality and cancer incidence data [6]. However, LSS is based on single acute exposure and still limited in its ability to precisely estimate site-specific cancer risks. Therefore, we acknowledge that the BEIR VII models need to be updated with other studies of chronic low-level radiation exposure with the longer duration follow-up.

The lifetime attributable risk (LAR) means the number of excess event incidences for an irradiated group compared with a non-radiation exposed group. The LAR risk projection model is defined as

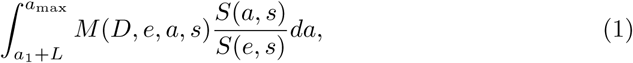

where *L* is the latency period for each cancer, *M* (*D, e, a, s*) is a risk model, and the ratio *S*(*a, s*)*/S*(*e, s*) is the conditional probability of a person alive and cancer-free at age-at exposure (*e*) to approach an attained age (*t*). We used cancer-free survival function after removing cancer incidence in the risk projection models [7]. In radiation epidemiology, LAR is the probability of premature cancer incidence attributable to radiation exposure in a representative member of the population [8–10] over their lifetime. World Health Organization (WHO) used LAR for Fukushima health risk assessment [11], and many other studies also have forecast radiation related cancer risk using LAR [3, 12–22].

The US National Cancer Institute ‘RadRAT’ program estimates lifetime risk of cancers induced by radiation exposure [23] based on BEIR VII [6], which estimated radiation related cancer risk transferred from Japanese (atomic bomb survivors) to US populations, and considered uncertainties related to risk model regression coefficients, e.g. ERR and EAR models (see [23] and [6] for more details), minimum latency, DDREF and radiation dose exposure distributions and population transfer weights. The ‘RadRAT’ program can be accessed for free at https://radiationcalculators.cancer.gov/radrat/ and is widely used to assess lifetime cancer risks attributable to radiation [20, 24, 25]. Although ‘RadRAT’ was designed to be easy and simple to use, it is difficult to flexibly use batch files for large data analyses or multiple cases, or ensure timely knowledge updates, e.g. baseline incidence rate in a specific population, life tables, etc. Thus, we developed an **LARisk** R package to calculate lifetime attributable risk for radiation related cancer built over the online RadRAT program. **LARisk** provides significantly enhanced flexibility by incorporating functions to project LAR for the batch file considering modified baseline incidence rate and/or population transfer weights, and DDREF application and a summary function based on average LAR for each case. In addition, **LARisk** uses the parametric bootstrap method to compute risk estimates and confidence intervals because the population distributions for uncertainties are known. The parametric bootstrap method is more powerful and provides small variances compared with the nonparametric bootstrap if the distribution assumption is known and the sample sizes are relatively small.

The remainder of this paper is organized as follows. Section **LAR models** describes the LAR risk models in detail, including the underlying ERR and EAR functions and discusses statistical uncertainties. Section **Illustration of LARisk package** discusses input information, output values, arguments, and result interpretation in the **LARisk** package; briefly describes **LARisk** options; Section **An example with chronic exposure rate** uses data examples to describe how to use **LARisk**. Finally, Section **Concluding remarks** summarizes the paper.

### LAR models

#### Risk model

Risk models for the **LARisk** package were based on those developed for the RadRAT program to estimate lifetime risk following low-dose radiation exposure. ERR (Excess Relative Risk) and EAR (Excess Absolute Risk) models were used for exposed individuals compared with unexposed individuals, based on a dose response function, *ρ*(*D*) with radiation dose *D* in Gray (Gy), and effect modification, ***ε*(X) = exp(*α***^***T***^ **X)** where **X** is a vector of several covariates including age at exposure, attained age, sex, etc. and *α* is a vector of regression coefficients. *ρ*(*D*) can be linear, linear-quadratic, quadratic, linear-threshold, nonparametric, etc. For example,

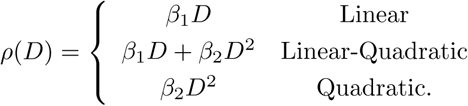

See [6], [1], [26], [27] and [28] for more details.

**LARisk** incorporates both ERR and EAR for a risk model (1) to compute LAR estimates of solid cancer and leukemia. The standard form of ERR or EAR models for solid cancer, except thyroid and breast, is defined as

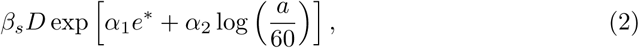

where the risk is assumed to be linearly related to the radiation dose, *a* is attained age, and *e* is age at exposure, which are considered to be effect modifications where

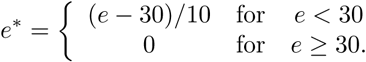

The form for thyroid and breast cancer is slightly different from the standard model. Thyroid only depends on age at exposure effect modification. The breast cancer model is defined as

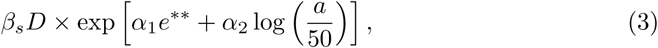

where *e*\*\* = (*e* − 25)*/*10, *α*_2_ is equal to 3.5 for *a* ≤ 50 and 1.0 elsewhere. Gallbladder, brain/CNS, and thyroid cancers only consider a risk model based on ERR to calculate LAR, and breast cancer utilizes a risk model based on only EAR. A standard form of a risk model for leukemia takes the model as

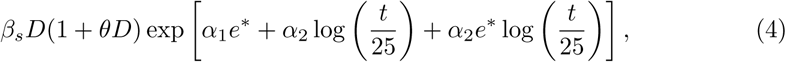

which is linear-quadratic in dose and age at exposure, and time since exposure (*t*) is considered as the effect modification. For chronic exposure, the risk model is linear with dose, i.e., *θ* = 0. A generalized nonlinear model, in particular a Poisson nonlinear model, is required to directly estimate the parameters [6, 26, 28, 29].

It is possible that LAR can be obtained from the multiplicative or additive model controlling weights. The multiplicative model assumes that radiation induced excess cancer risk proportionally increases with baseline risk as

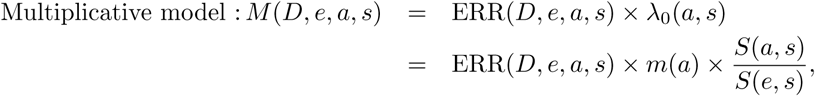

where *λ*_0_(*a, s*) is the excess risk for non-exposed group. On the other hand, the additive model assumes that radiation induced excess cancer risk increases independently of baseline risk as

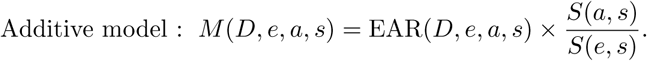

Both LAR_ERR_ and LAR_EAR_, are LAR components based on ERR and EAR models, respectively, defined as

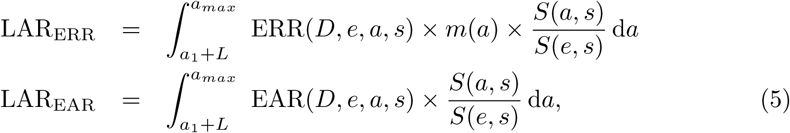

where *m*(*a*) is population cancer incidence rate, and *S*(*a, s*)*/S*(*e, s*) is the probability of surviving to age *a*, conditional on survival to age *e*, given sex *s*. Thus, the LAR model in the **LARisk** package is computed as

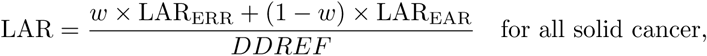

and

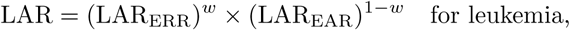

where *w* is a population transfer weight.

### Uncertainty assessment

Uncertainties including statistical uncertainties must be considered when calculating LAR, since we use probability distribution functions based on radiation epidemiology and Monte Carlo (MC) simulation. The uncertainties are as follows.

1. risk model coefficients,
2. dose and dose-rate effectiveness factor,
3. minimum latency,
4. risk population transfer between populations (e.g. Japan to US),
5. radiation doses, and
6. baseline incidence rate.

Uncertainties 1–4 including statistical uncertainties have the most critical impact to assess lifetime cancer risk attributable to radiation, whereas uncertainties 5 and 6 cause LAR variability regardless of the model. The baseline incidence rate in particular (uncertainty 6) differs with population and year, generating different incidence rate distributions by age.

To calculate radiation induced future cancer risk, we substitute the formula (2) into (5) for solid cancer as

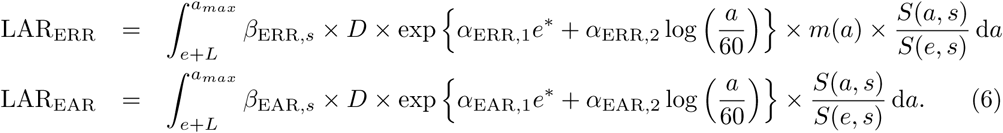

The regression coefficients, *β*_ERR,*s*_, *β*_EAR,*s*_, *α*_ERR,1_, *α*_EAR,1_, *α*_ERR,2_, and *α*_EAR,1_, were determined from radiation epidemiologic research with atomic bomb (A-bomb) survivors at RERF, which has conducted health related research among A-bomb survivors in Hiroshima and Nagasaki, Japan, for more than 70 years. Tables 1 and 2 show the regression coefficients for solid cancers and leukemia, respectively. For leukemia, we replace the formula (2) by (4) to derive a similar expression to the formula (6).

**Table 1.** The regression coefficients of all radiation related solid cancer risk models [6, 23].

| **ERR regression coefficients for solid cancers** |  |  |  |  |
| --- | --- | --- | --- | --- |
| Cancer site | $\beta_{Male}$ | $\beta_{Female}$ | $\alpha_1$ | $\alpha_2$ |
| Oral | 0.23 (<0.0–0.66) | 0.53 (0.13–1.24) | –0.30 | –1.40 |
| Esophagus | 0.51 (<0.0–1.13) | 0.82 (<0.0–3.10) | –0.30 | –1.40 |
| Stomach | 0.21 (0.11–0.40) | 0.48 (0.31–0.73) | –0.30 | –1.40 |
| Colon | 0.63 (0.37–1.10) | 0.43 (0.19–0.96) | –0.30 | –1.40 |
| Rectum | 0.12 (<0.0–0.38) | 0.12 (<0.0–0.38) | –0.30 | –1.40 |
| Gallbladder | –0.018 (<0.0–0.29) | –0.018 (<0.0–0.29) | –0.30 | –1.40 |
| Pancreas | 0.36 (<0.0–0.88) | 0.36 (<0.0–0.88) | –0.30 | –1.40 |
| Liver | 0.32 (0.16–0.64) | 0.32 (0.10–1.00) | –0.30 | –1.40 |
| Lung | 0.32 (0.15–0.70) | 1.4 (0.94–2.10) | –0.30 | –1.40 |
| Breast | – | – | – | – |
| Ovary | – | 0.38 (0.10–1.40) | –0.30 | –1.40 |
| Uterus | – | 0.05 (<0.0–0.22) | –0.30 | –1.40 |
| Prostate | 0.12 (<0.0–0.69) | – | –0.30 | –1.40 |
| Bladder | 0.50 (0.18–1.40) | 1.65 (0.69–4.00) | –0.30 | –1.40 |
| Kidney | 0.34 (<0.0–1.00) | 0.34 (<0.0–1.00) | –0.30 | –1.40 |
| Brain/CNS | 0.71 (0.26–1.34) | 0.24 (0.09–0.47) | –0.30 | –1.40 |
| Thyroid | 0.53 (0.14–2.00) | 1.05 (0.28–3.90) | –0.83 | 0.00 |
| Remainder | 0.84 (0.45–1.69) | 0.80 (0.33–1.93) | –0.30 | –1.40 |

| **EAR regression coefficients for solid cancers** |  |  |  |  |
| --- | --- | --- | --- | --- |
| Cancer site | $\beta_{Male}$ | $\beta_{Female}$ | $\alpha_1$ | $\alpha_2$ |
| Oral | 0.44 (0.08–1.10) | 0.29 (0.06–0.66) | –0.41 | 0.50 |
| Oesophagus | 0.88 (0.11–2.10) | 0.14 (<0.0–0.63) | –0.41 | 2.80 |
| Stomach | 4.90 (2.70–8.90) | 4.90 (3.20–7.30) | –0.41 | 2.80 |
| Colon | 3.20 (1.80–5.60) | 1.60 (0.80–3.20) | –0.41 | 2.80 |
| Rectum | 0.34 (0.09–1.10) | 0.34 (0.09–1.10) | –0.41 | 2.80 |
| Gallbladder | – | – | – | – |
| Pancreas | 0.49 (0.09–1.10) | 0.49 (0.09–1.10) | –0.41 | 2.80 |
| Liver | 2.20 (0.90–5.30) | 1.00 (0.40–2.50) | –0.41 | 4.10 |
| Lung | 2.30 (1.10–5.00) | 3.40 (2.30–4.90) | –0.41 | 5.20 |
| Breast | – | 10.00 (7.00–14.20) | –0.50 | 3.50, 1.00 |
| Ovary | – | 0.70 (0.20–2.10) | –0.41 | 2.80 |
| Uterus | – | 1.20 (<0.0–2.60) | –0.41 | 2.80 |
| Prostate | 0.11 (<0.0–1.0) | – | –0.41 | 2.80 |
| Bladder | 1.20 (0.40–3.70) | 0.75 (0.30–1.70) | –0.41 | 6.00 |
| Kidney | 0.31 (0.08–0.68) | 0.31 (0.08–0.68) | –0.41 | 2.80 |
| Brain/CNS | – | – | – | – |
| Thyroid | – | – | – | – |
| Remainder | 2.73 (1.55–4.82) | 1.06 (0.49–2.28) | –0.41 | 2.80 |

Note that the values in the parentheses represent 95% confidence intervals for each regression coefficient.

**Table 2.** Regression coefficients for radiation related leukemia risk models [6, 23]

| **ERR and EAR models for leukemia** |  |  |  |
| --- | --- | --- | --- |
| | $\beta_{Male}$ | $\beta_{Female}$ | $\alpha_1$ |
| ERR | 1.10 (0.10–2.60) | 1.20 (0.10–2.90) | −0.40 (−0.78 to 0.00) |
| EAR | 1.62 (0.10–3.60) | 0.93 (0.10–2.00) | 0.29 (0.00–0.62) |
| | $\alpha_2$ | $\alpha_3$ | $\theta$ |
| ERR | −0.48 (−1.10 to 0.20) | 0.42 (0.00–0.96) | 0.87 (0.16–15.00) |
| EAR | 0.00 | 0.56 (0.31–0.85) | 0.88 (0.16–15.00) |

Note that the values in the parentheses represent 95% confidence intervals for each regression coefficient.

The uncertainties of risk model coefficients for solid cancers focus on sex-specific main effect parameters, whereas other parameters are specific constants derived from A-bomb survivor epidemiology studies. Table 1 shows uncertainties for ERR and EAR model parameters. Dose coefficient, *β*, for stomach, colon, liver, lung, breast, ovary, bladder, and thyroid cancer is assumed to follow log-normal distribution, hence regression coefficient distributions in LAR models for prostate and uterus cancer are also considered to be normally distributed. Uncertainty in *β* is described by cumulative distribution functions (CDFs) for oral, oesophagus, gallbladder, pancreas, rectum, kidney, and brain/CNS cancers. *α*_2_ of breast cancer has two values. See the formula 3. In contrast to solid cancer, uncertainty for leukemia is associated with five parameters. Since the covariance matrix for regression coefficients in the leukemia LAR model should be considered, we calculate an approximate estimator for lifetime risk variance using the delta method following the BEIR VII committee [6].

The dose and dose-rate effectiveness factor (DDREF) combines two concepts of a low-dose effectiveness (LDEF) and a dose-rate effectiveness factor (DREF) [6, 30, 31]. LDEF means a ratio of the slope of a linear-quadratic dose-response function for acute exposure and the slope of the linear term of the same model, whereas DREF is computed when an acute exposure is divided and the number of fractions is large enough. ICRP suggested to utilize a DDREF of 2 to reduce solid cancer rates obtained from moderate-to-high acute dose studies for low-dose or low-dose rate exposures [31, 32]. The DDREF distribution is considered as a lognormal with geometric mean= 1.5 and standard deviation= 1.35 [23]. DDREF is always applied for chronic exposure without dose-dependence. In acute exposures, DDREF is applied by comparing dose limit, *D*_*L*_, and exposure dose. If dose *< D*_*L*_ (generated 30 − 200 mGy using log-uniform distribution), DDREF is considered as used. DDREF is not applied for leukemia, because the uncertainty associated with low-dose risks is modeled using a linear term.

Latency uncertainty for radiation-induced cancers should be considered, because each person has cancer different development time after radiation exposure. Latency distribution is commonly represented by an S-shaped curve with the parameter in the center of the S-shaped function generated from the triangular distribution for solid cancer, thyroid and leukemia as *T* (5, 7.5, 10), *T* (3, 5, 7) and *T* (2, 2.25, 2.5), respectively. An uncertain latency adjustment is between 4 and 11 years after radiation exposure for solid cancer, 2.5 and 7.6 years for thyroid cancer, and 0.4 and 4.1 years for leukemia [33].

We must also consider effect of transferring the risk from Japanese population to population of interest. Because parameters of risk model coefficients are commonly obtained using the Japanese A-bomb survivors cohort, the effect is explained by the weighted mean of LAR values based on ERR and EAR model estimates. Initially we use the RadRAT weight to transfer the risk from Japanese to US populations. Thus, the weight *w* = 0.7 is assigned to the ERR model and the remaining (1 − *w* = 0.3) weight to the EAR model for most cancers. Lung cancer weight is reversed, and only the EAR model for breast is used. For thyroid, gallbladder, and brain/CNS, only ERR is used for calculation of LAR.

Radiation dose uncertainty can be applied to a fixed value or various probability distributions, including lognormal, normal, triangular, log-triangular, uniform, and log-uniform.

The baseline incidence rate depends on population information.

### Parametric bootstrap

#### Algorithm 1: Computation of parametric bootstrap confidence intervals for risk estimates

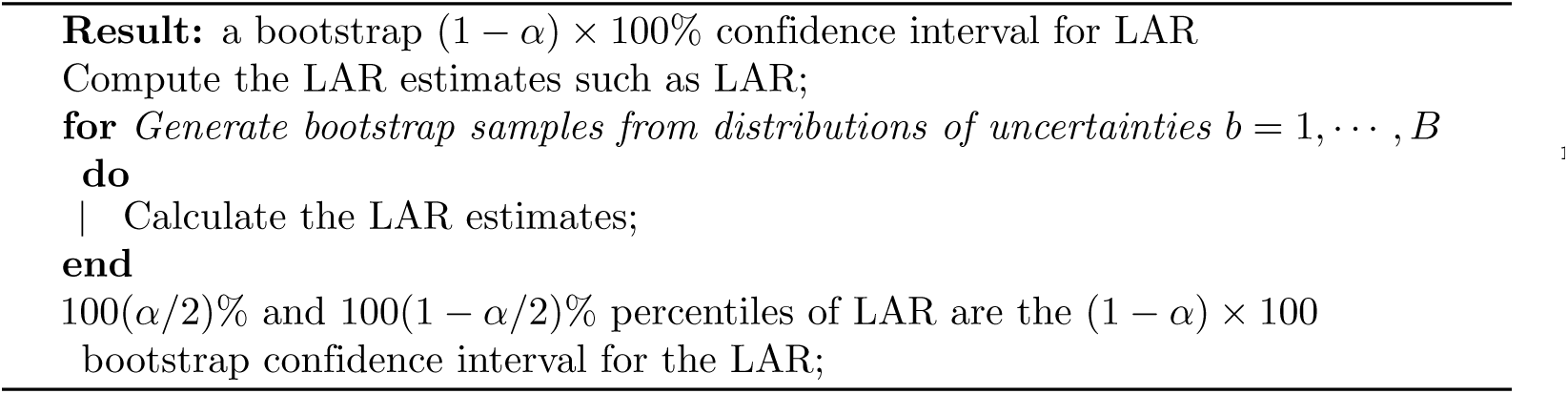

Since uncertainties’ distributions for estimation of LAR are already known, we apply the parametric bootstrap to compute the confidence intervals for all risk estimates. The parametric bootstrap needs distribution assumption for generation and the other procedure is identical to the general nonparametric bootstrap. The parametric bootstrap is more powerful, in that the method provides small variances compared with the nonparametric bootstrap if the distribution is assumed to be right and the sample sizes are relatively small. The parametric bootstrap algorithm is provided in Algorithm 1.

### Illustration of LARisk package

#### Main arguments

The **LARisk** package has 3 main functions for estimating lifetime attributable risks such as **LAR, LAR_batch** and **LAR_group**. The primary argument for the **LAR** function is as follows.

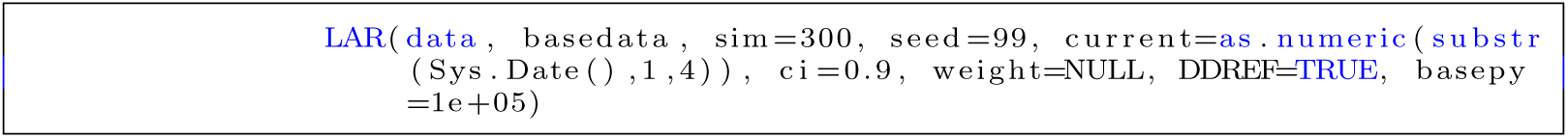

The argument ‘data’ should include some prerequisite information such as sex, and birth year(s), exposure year, distributions of exposure dose, radiation dose, cancer sites, and exposure rate, shown as follows.

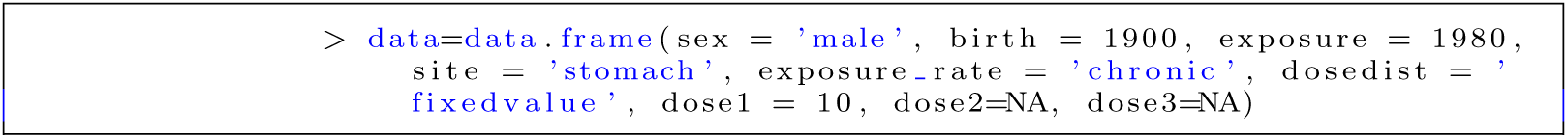

For ‘site’, we provide the irradiated organ or cancer site. In ‘dosedist’, ‘dose1’, ‘dose2’, and ‘dose3’ arguments, we insert the distribution of the exposure dose such as ‘fixedvalue’, ‘lognormal’, ‘normal’, ‘triangular’, ‘logtriangular’, ‘uniform’ or ‘loguniform’ with appropriate parameters for each distribution. For instance, if the exposure dose has a normal distribution with mean of 2.3 and standard deviation of 0.8, we set ‘dose1=2.3’, ‘dose2=0.8’, and ‘dose3=NA’. If the dose has fixed value of 3.2, ‘dose1=3.2’, ‘dose2=NA’ and ‘dose3=NA’ should be employed. If new population information should be considered, lifetime and cancer incidence rate tables are employed in a ‘basedata’ argument as follows.

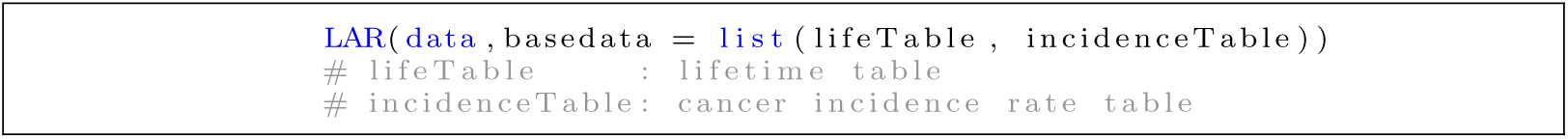

**LARisk** has default for these tables, which were made in 2010 and 2018, South Korea such as ‘life2010’, ‘incid2010’, ‘life2018’ and ‘incid2018’. Lifetime and cancer incidence rate tables should also have the specified format as follows.

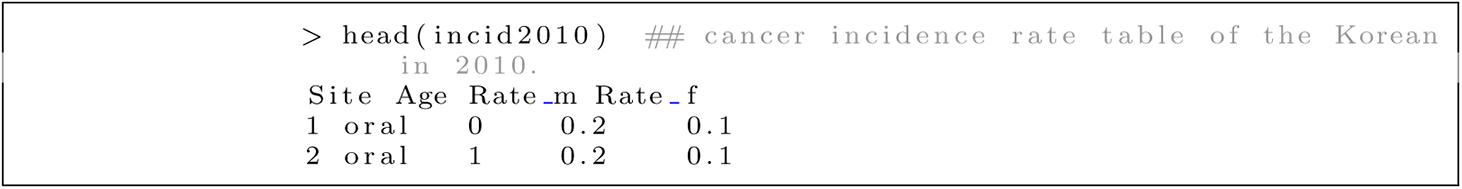

Also, the cancer incidence rate table consists of ‘Site’, ‘Age’, ‘Rate_m’, and ‘Rate_f’ where ‘Rate_m’ and ‘Rate_f’ are incidence rates of each cancer site according to male and female, respectively.

The argument ‘weight’ is employed to compute LAR values through the weighted average of LAR values based on ERR and EAR models. For example, if the ‘weight’ for LAR based on between EAR and ERR models for stomach cancer is set to 0.5, run the below code.

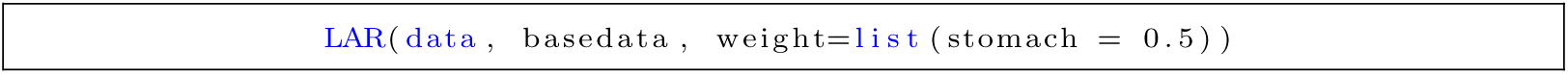

**LARisk** sets the default weights to 0.7 for most cancers, 0.3 for lung cancer, 0.0 for breast and1.0 for thyroid for LAR based on EER (see Table 3).

**Table 3.** Weights for each cancer site.

| Cancer site | ERR | EAR | weight |
| --- | --- | --- | --- |
| Most cancer | 0.7 | 0.3 | 0.7 |
| Lung | 0.3 | 0.7 | 0.3 |
| Breast | 0.0 | 1.0 | 0.0 |
| Thyroid | 1.0 | 0.0 | 1.0 |
| Gallbladder | 1.0 | 0.0 | 1.0 |
| Brain/CNS | 1.0 | 0.0 | 1.0 |

Dose and dose-rate effectiveness factor (DDREF) is the logical option to select whether or not to consider ‘DDREF’ for computing LAR values. The value ‘DDREF’ is to modify the effect of exposure to low-dose. The value of ‘DDREF’ is considered differently as per exposure rate. However, if the site is leukemia, DDREF dose not apply even if DDREF = TRUE.

### Estimating LAR values using LAR, LAR_batch and LAR_group

To describe how to use functions installed in the **LARisk** package, we use the toy data ‘nuclear’, also installed in **LARisk**, assuming scenario of the data in which everyone is exposed to radiation at the same time in 2011. The data includes 20 people for 10 males and 10 females, and the radiation exposed age has ranges from 3 to 81 years with acute exposed rate and fixed dose distribution.

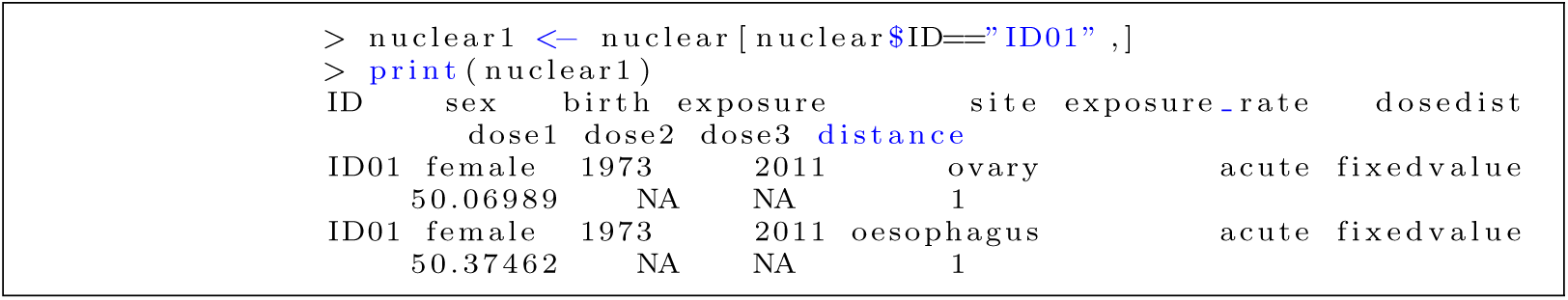

**LAR, LAR_batch**, and **LAR_group** functions provide 3 main estimated values such as lifetime risk, future risk and lifetime baseline risk. Among them, ‘LAR’ and ‘F_LAR’ in Table 4 have mean values and confidence limits (lower and upper) for each cancer site, solid cancer and total (solid cancer+leukemia), respectively. See Table 4 for more details.

**Table 4.** Values in the LARisk package.

| Value | Description |
| --- | --- |
| LAR | Lifetime attributable risk (Cancer incidence probability per 100,000 persons after exposed year) |
| F_LAR | Future LAR (LAR after current year) |
| LBR | Lifetime baseline risk (Cumulative baseline probability per 100,000 persons of having cancer over the maximum lifetime without radiation exposure after exposed year) |
| BFR | Baseline future risk (Cumulative baseline probability per 100,000 persons of having cancer over the maximum lifetime without radiation exposure after current year) |
| LFR | Lifetime fractional risk (=LAR/LBR) |
| TFR | Total future risk (= F_LAR + BFR) |

The **LAR** function prints the total LAR, future LAR, baseline future risk, and total future risk. If you want to obtain more detailed result, you can use the ‘summary’ function. The ‘summary’ function provides the person’s gender and year of birth, and risks by cancer type, confidence level, and current year. In the result, the ‘LAR’ tab includes site-specific LAR, LBR, and LFR, whereas the ‘Future LAR’ tab contains site-specific future LAR, BFR, and TFR.

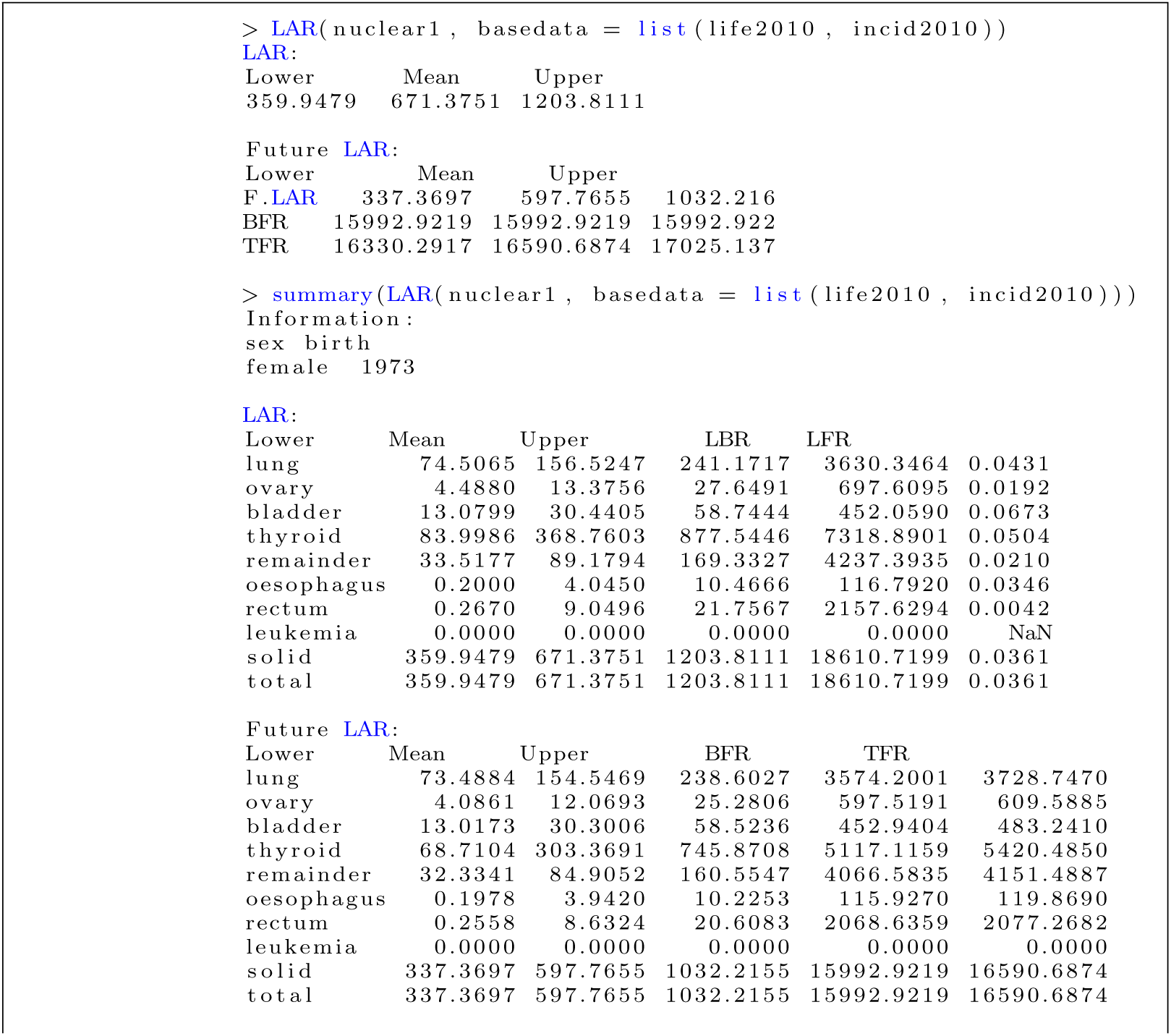
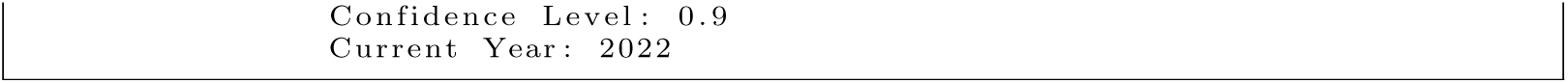

If you compute LAR values for many individuals, you can use the **LAR_batch** function. Unlike **LAR, LAR_batch** calculates each person’ risk after reading multiple people’s data at once. For example, suppose that we want to calculate the LAR values in the toy data ‘nuclear’ with person “ID”. The sums of LAR values of all cancers for 3 observations are computed as follows. Here the argument ‘max.id’ means the number of individuals that should be printed.

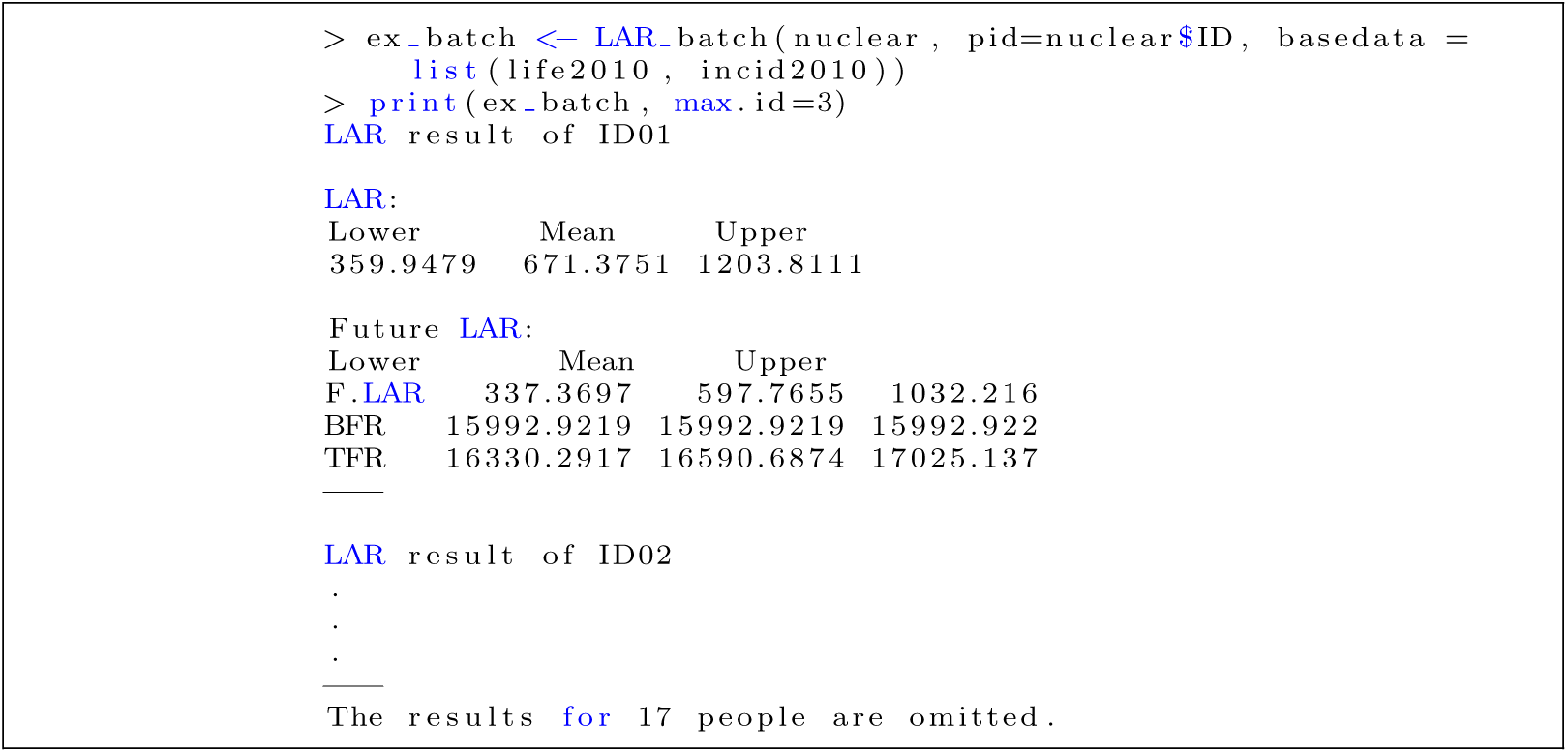

Similarly, using the ‘summary’ function, you can get more detailed results for each cancer site as follows.

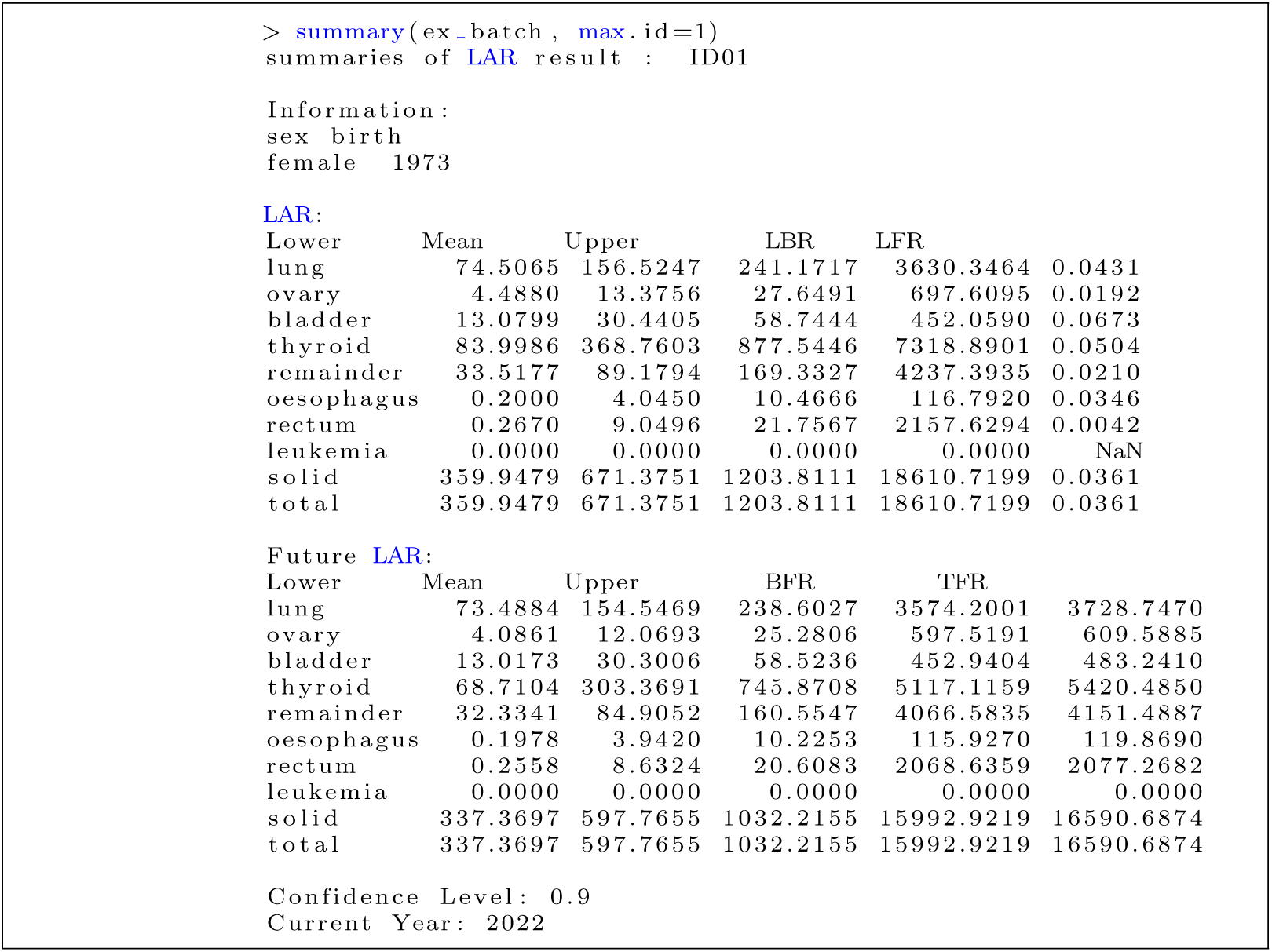

The **LAR_group** function averages estimated LAR values by group, which provides grouped LAR, grouped future LAR, and grouped baseline risk values for each group. The average LAR values in the toy data ‘nuclear’ can be computed by ‘distance’ as follows.

| Lower | Mean | Upper | LBR | LFR |  |
| --- | --- | --- | --- | --- | --- |
| stomach | 50.9850 | 66.2373 | 87.8936 | 2926.1714 | 0.0226 |
| colon | 43.4301 | 56.1274 | 72.3353 | 2160.0832 | 0.0260 |
| liver | 7.0657 | 10.4933 | 15.2987 | 808.9772 | 0.0130 |
| lung | 39.1538 | 53.4791 | 70.6998 | 3097.9930 | 0.0173 |
| breast | 2.8636 | 4.2821 | 6.0646 | 331.2576 | 0.0129 |
| ovary | 0.3206 | 0.9554 | 1.9749 | 49.8292 | 0.0192 |
| uterus | 0.0216 | 0.1491 | 0.3288 | 68.4665 | 0.0022 |
| prostate | -15.4917 | 6.2743 | 27.6037 | 960.8121 | 0.0065 |
| bladder | 10.5122 | 16.1598 | 23.8353 | 430.7553 | 0.0375 |
| brain/cns | 0.7414 | 1.3918 | 2.2998 | 46.3592 | 0.0300 |
| thyroid | 67.7618 | 185.2396 | 350.2187 | 1648.0018 | 0.1124 |
| remainder | 34.5468 | 56.8793 | 88.6948 | 912.3992 | 0.0623 |
| oral | 0.3038 | 1.3365 | 2.7663 | 105.2675 | 0.0127 |
| oesophagus | 2.7255 | 5.3313 | 8.1798 | 208.5186 | 0.0256 |
| rectum | 0.3667 | 1.3777 | 2.5946 | 618.5623 | 0.0022 |
| gallbladder | -6.2708 | -0.9336 | 4.2742 | 420.9955 | -0.0022 |
| pancreas | 3.4377 | 7.4064 | 11.9408 | 493.6795 | 0.0150 |
| kidney | 1.0530 | 3.2226 | 6.5984 | 203.1165 | 0.0159 |
| leukemia | 0.0713 | 0.2012 | 0.5677 | 19.9927 | 0.0101 |
| solid | 339.2210 | 475.4092 | 659.3305 | 15491.2456 | 0.0307 |
| total | 339.4012 | 475.6104 | 659.5318 | 15511.2384 | 0.0307 |

Future LAR:
| Lower | Mean | Upper | BFR | TFR |  |
| --- | --- | --- | --- | --- | --- |
| stomach | 48.2294 | 63.3542 | 84.7154 | 2490.0371 | 2553.3913 |
| colon | 40.9639 | 53.4541 | 69.9636 | 1902.1381 | 1955.5921 |
| liver | 6.0418 | 9.0587 | 13.2437 | 571.8794 | 580.9381 |
| lung | 38.4091 | 52.5642 | 69.9525 | 2877.2243 | 2929.7885 |
| breast | 2.3678 | 3.5644 | 5.0447 | 157.2215 | 160.7859 |
| ovary | 0.2919 | 0.8621 | 1.8058 | 42.6799 | 43.5420 |
| uterus | 0.0203 | 0.1258 | 0.2655 | 41.5060 | 41.6318 |
| prostate | -15.5887 | 6.2226 | 27.5255 | 881.1613 | 887.3839 |
| bladder | 10.3276 | 16.0169 | 23.7049 | 387.4721 | 403.4890 |
| brain/cns | 0.6220 | 1.1373 | 1.8552 | 35.4464 | 36.5837 |
| thyroid | 60.0224 | 170.7748 | 330.0385 | 1233.1081 | 1403.8830 |
| remainder | 33.7594 | 54.9135 | 84.8616 | 837.8480 | 892.7615 |
| oral | 0.2668 | 1.2582 | 2.6651 | 87.9061 | 89.1643 |
| oesophagus | 2.4342 | 4.4985 | 6.7130 | 162.4755 | 166.9740 |
| rectum | 0.3337 | 1.2961 | 2.4649 | 524.0167 | 525.3128 |
| gallbladder | -6.0139 | -0.8791 | 4.1981 | 361.3716 | 360.4925 |
| pancreas | 3.2450 | 7.1431 | 11.5463 | 453.7292 | 460.8723 |
| kidney | 0.9721 | 3.1387 | 6.5336 | 183.9500 | 187.0887 |
| leukemia | 0.0827 | 0.1485 | 0.2667 | 13.7139 | 13.8624 |
| solid | 318.3267 | 448.5040 | 621.6234 | 13231.1713 | 13679.6753 |
| total | 318.4752 | 448.6524 | 621.7718 | 13244.8852 | 13693.5376 |

``` Confidence Level: 0.9 Current Year: 2022 ```
``` summaries of LAR result : Group 2 ```
``` Group Information: sex count birth female 12 1987.417 male 5 1956.000 ```

### Saving outputs

**LARisk** includes a function **write_LAR** which writes outputs from **LAR, LAR_batch**, and **LAR_group** into a CSV file.

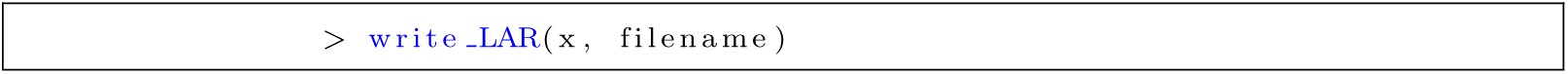

The format of the saved file is illustrated in Table 5.

**Table 5.** Format to save outputs using a write_LAR function.

|  | Lower | Mean | Upper | F.Lower | F.Mean | F.Upper | LBR | BFR | LFR | TFR |
| --- | --- | --- | --- | --- | --- | --- | --- | --- | --- | --- |
| site-name | . | . | . | . | . | . | . | . | . | . |
| solid | . | . | . | . | . | . | . | . | . | . |
| total | . | . | . | . | . | . | . | . | . | . |

### An example for chronic exposure rate with a toy data ‘organ’

In this section, we consider a toy data ‘organ’ which consists of the 20 radiation exposed people over various periods with 14 males and 6 females having the job information ‘occup’. The data is shown as follows.

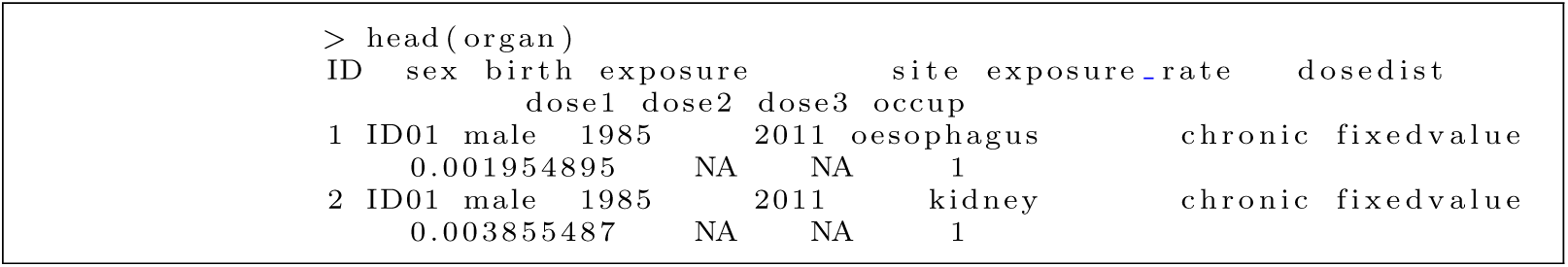

Suppose that we want to calculate LAR values with the current year=2021 using the Korean population in 2018. First, we compute LAR values with ‘ID01’ as follows.

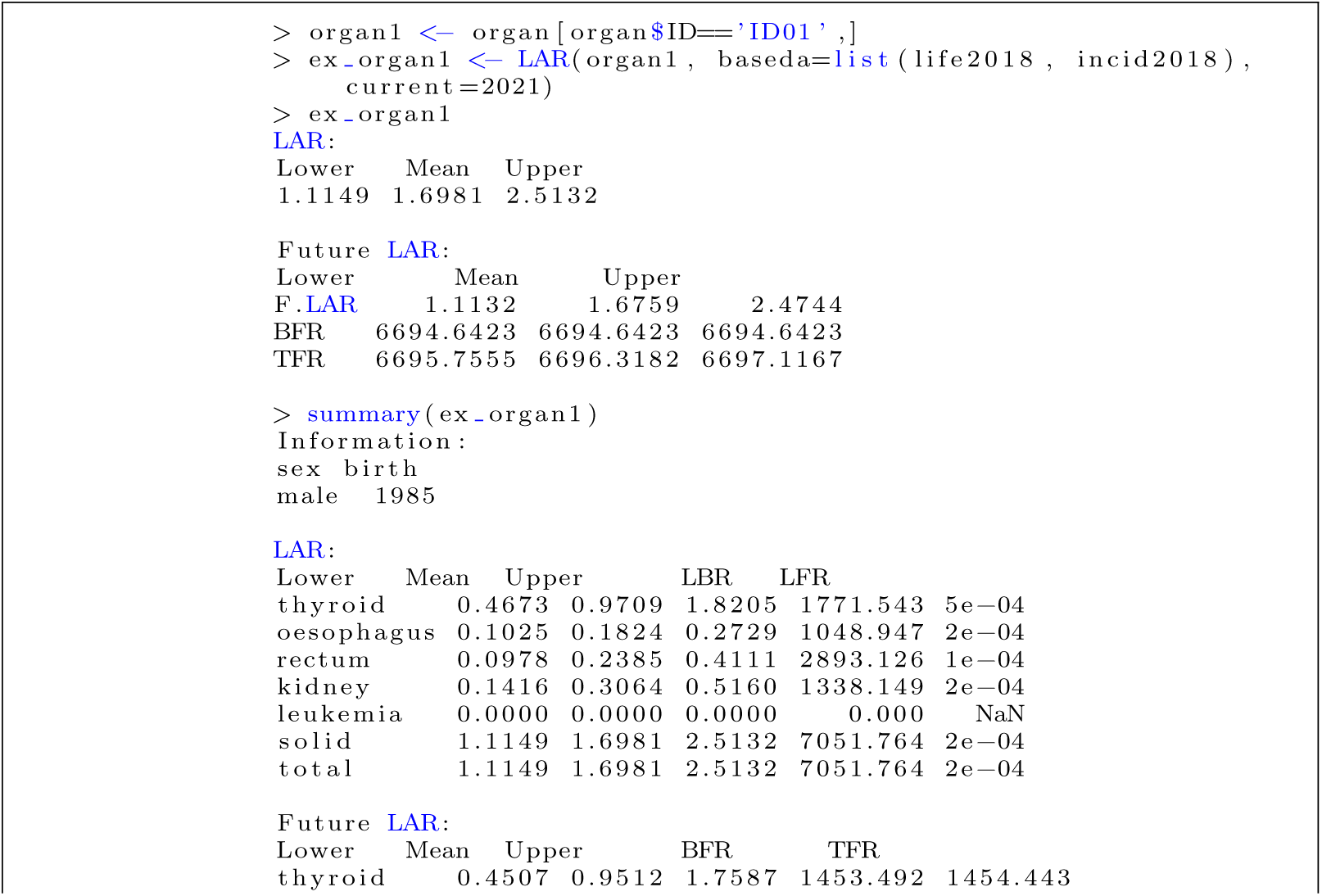
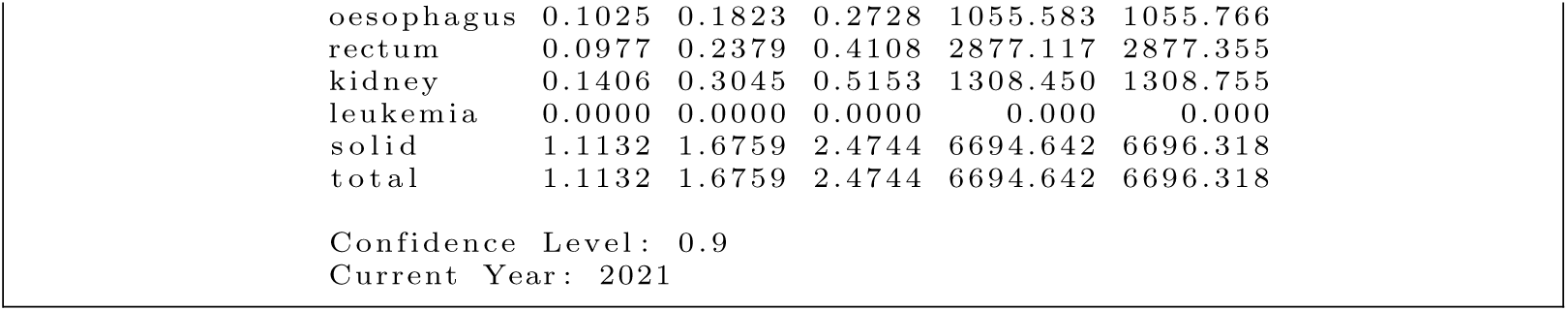

The estimated total LAR values of the person ‘ID01’ is 1.6981 with the 90% confidence interval (1.1149, 2.5132) and the future risk is 1.6759 with the 90% confidence interval (1.1132, 2.4744). The person ‘ID01’ is a man born in 1985. ‘ID01’ was exposed to thyroid, oesophagus, ‘rectum’, and kidney except for leukemia. Thus, The LAR values of leukemia are all 0.

We also compute LAR values by group - male and female using **LAR_group** as follows.

| Lower | Mean | Upper | LBR | LFR |  |
| --- | --- | --- | --- | --- | --- |
| colon | 0.6832 | 1.1392 | 1.7064 | 1080.1190 | 0.0011 |
| lung | 2.3739 | 2.8990 | 3.5941 | 1353.2750 | 0.0021 |
| uterus | 0.1447 | 0.2874 | 0.4696 | 338.1363 | 0.0008 |
| bladder | 0.9560 | 1.3026 | 1.7539 | 155.9481 | 0.0084 |
| remainder | 2.9905 | 4.5565 | 6.7692 | 1875.6871 | 0.0024 |
| oral | 0.1509 | 0.2311 | 0.3413 | 130.7238 | 0.0018 |
| oesophagus | 0.0203 | 0.0768 | 0.1583 | 18.0691 | 0.0043 |
| rectum | 0.2375 | 0.4315 | 0.6480 | 873.1753 | 0.0005 |
| gallbladder | -0.1736 | -0.0274 | 0.1067 | 290.7825 | -0.0001 |
| pancreas | 0.0661 | 0.1089 | 0.1608 | 283.0072 | 0.0004 |
| leukemia | 0.0792 | 0.1800 | 0.4092 | 81.1517 | 0.0022 |
| solid | 9.3445 | 11.0056 | 13.3345 | 6398.9234 | 0.0017 |
| total | 9.5265 | 11.1856 | 13.5145 | 6480.0751 | 0.0017 |

Future LAR:
| Lower | Mean | Upper | BFR | TFR |  |
| --- | --- | --- | --- | --- | --- |
| colon | 0.6658 | 1.0964 | 1.6314 | 1051.6541 | 1052.7505 |
| lung | 2.3673 | 2.8906 | 3.5837 | 1352.5231 | 1355.4137 |
| uterus | 0.1423 | 0.2825 | 0.4686 | 296.3270 | 296.6095 |
| bladder | 0.9503 | 1.2916 | 1.7359 | 154.4180 | 155.7096 |
| remainder | 2.8942 | 4.4153 | 6.5409 | 1792.3971 | 1796.8124 |
| oral | 0.1444 | 0.2181 | 0.3177 | 119.1196 | 119.3377 |
| oesophagus | 0.0201 | 0.0762 | 0.1568 | 18.0923 | 18.1685 |
| rectum | 0.2346 | 0.4186 | 0.6327 | 835.6204 | 836.0390 |
| gallbladder | -0.1735 | -0.0274 | 0.1066 | 292.4045 | 292.3771 |
| pancreas | 0.0653 | 0.1079 | 0.1591 | 281.1836 | 281.2915 |
| leukemia | 0.0944 | 0.1773 | 0.3603 | 77.5662 | 77.7435 |
| solid | 9.1382 | 10.7697 | 12.9982 | 6193.7399 | 6204.5095 |
| total | 9.3172 | 10.9470 | 13.1770 | 6271.3060 | 6282.2530 |

Confidence Level: 0.9 Current Year: 2021
``` summaries of LAR result : Group male ```
``` Confidence Level: 0.9 Current Year: 2021 ```

The above results illustrate that the estimated average LAR values (90% confidence intervals) for female and male are 11.1856(9.5265, 13.5145) and 27.1674(23.8700, 28.7939), respectively. **LAR_group** can consider two group variables such as gender and occupation in this toy ‘organ’ data as follows.

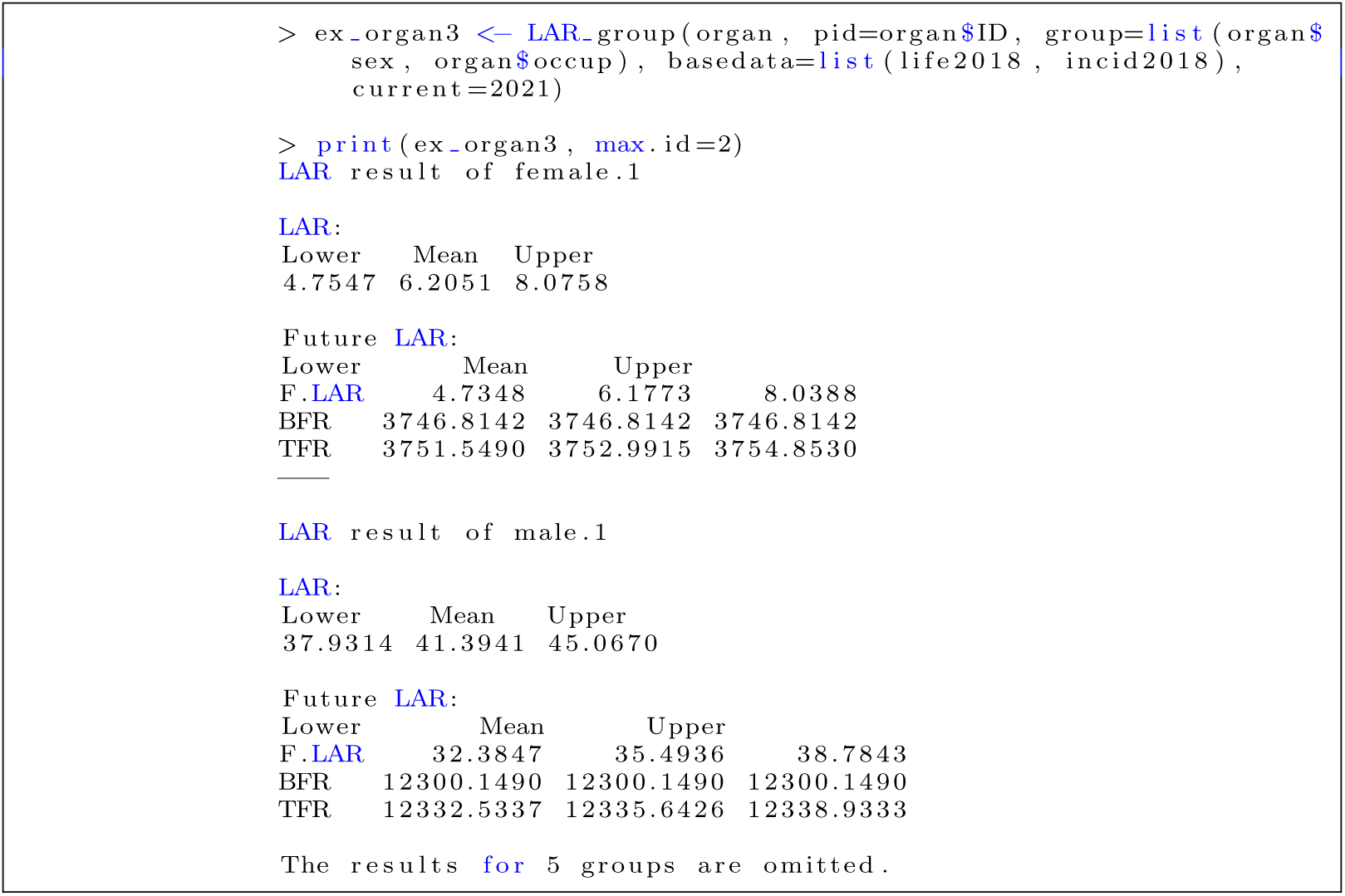

### Concluding remarks

We proposed the **LARisk** package built in R to calculate projected and/or lifetime radiation related cancer risks after exposure. We described and considered the various uncertainties in the lifetime risk calculations to construct this package. Three main flexible and useful functions were developed, and many arguments/options are available to promptly update new knowledge from radiation epidemiology and related fields. Of particular interest is the ability to input population information, replacing cancer incidence rates and survival functions for the population of interest and observed year to be used for lifetime risk calculation. This greatly simplifies to employ new baseline incidence rate and population survival rate data for a given year. Detailed description of the arguments/options and related data formats, etc. are provided in the **LARisk** vignette (https://cran.r-project.org/web/packages/LARisk/vignettes/LARisk-vignette.html).

**LARisk** is an effective and simple method to calculate excess lifetime risk from radiation exposure, and can be applied to any population of interest.

## Data Availability

Toy data sets in LARisk R-package

https://cran.r-project.org/web/packages/LARisk/index.html

